# Apraxia as a Predictor of Post-Stroke Recovery: Insights from the Birmingham Cognitive Screening Program

**DOI:** 10.1101/2025.01.25.25321111

**Authors:** Elisabeth Rounis, Siddharth Ramanan, Wai-Ling Bickerton, Nele Demeyere, Matthew A. Lambon Ralph

## Abstract

Our study highlights cognitive measures underlying post-stroke recovery using the ‘Birmingham Cognitive Screening’ (BCOS) program—the only dataset with comprehensive assessments of cognition, praxis, and stroke outcomes. We analysed 256 stroke patients tested for cognitive deficits, which uniquely included apraxia and the Barthel Index for daily living activities at subacute and chronic stages. Using step-wise multivariate linear regression, we identified significant predictive factors—praxis, language, orientation, and baseline independence—linked to improved daily activities, and validated our model with 4-fold cross-validation. This research underscores the vital role of specific cognitive measures including apraxia as key cognitive predictors for stroke recovery.

## Introduction

Stroke is the global leading cause of morbidity. Despite improvements in mortality rates with advanced acute stroke treatments such as thrombolysis and thrombectomy, two-thirds of stroke survivors still leave UK hospitals with a disability(1). The imperative to assess for cognitive deficits is underscored by emerging evidence suggesting their potential to forecast post-stroke recovery. Among these deficits, limb apraxia emerges as a consequence of stroke, which is linked to prolonged disability and dependence(2,3). This disorder impedes skilled actions, manifesting in challenges with everyday tasks such as combing hair, dressing, and preparing a cup of coffee. The intricate demands of neuropsychological testing coupled with the diverse manifestations of apraxia complicate the systematic assessment of patients’ deficits in clinical settings. Consequently, formulating targeted neuro-rehabilitative or compensatory strategies becomes challenging, with patients frequently leaving acute stroke units uninformed about their condition and lacking suitable management strategies.

Few studies encompass a comprehensive array of neuropsychological assessments necessary to identify apraxia and other cognitive deficits post-stroke, aligning them with stroke outcome measures. Moreover, there are very few longitudinal studies investigating how complex neuropsychological deficits may recover over time; and/or whether they lead to worse outcomes. Cognitive difficulties following a stroke are typically evaluated through either brief yet somewhat constrained general tests or exhaustive but time-consuming assessments that focus on specific domains such as language, memory, neglect and praxis that are difficult to deliver in the acute setting.

Addressing these challenges, we leverage the ‘Birmingham Cognitive Screening’ (BCoS) study dataset, which uniquely comprised both ‘broad’ and ‘shallow’ neuropsychological testing of multiple cognitive abilities, sampling these briefly to be time(4). BCoS has been validated as an efficient screen to identify patients with different forms of praxic deficit designed to be inclusive for patients with aphasia and/or spatial neglect(4).

We analysed data from 256 patients tested on this screen both at a sub-acute stage of stroke (<1 month) at the chronic post-stroke stages (>9 months). All patients were tested on a comprehensive set of cognitive tasks and activities of daily activities on the Barthel Index (BI-ADL) at both time points. We applied multivariate prediction models using step-wise regressions to identify measures at the early subacute stages that best predicted a change in BI-ADL at the chronic stage, and internally validated this model using internal 4-fold cross-validation.

## Methods

### Participants & Measures

This study utilised the dataset from stroke participants who took part in the ‘BCOS’ cohort study(4). Demographics and scores for patients are outlined in **Table 1**. A total of 256 patients were included and tested at two time points (sub-acute: <1month; chronic: >9month) on 34 cognitive measures, five of which related to praxis. Praxis testing was based on the BCOS time-efficient version of a validated apraxia battery that maintain sensitivity to both mild and severe impairments(5). Its domains included (i) pantomime to auditory/written words input adapted from the Florida Apraxia Task(6) (gesture production); (ii) forced choice recognition of pantomime adapted from Peigneux et al. 2000 (7) (gesture recognition); (iii) imitation of meaningless gestures adapted from De Renzi et al. 1980 (8) (meaningless imitation); (iv) a multi-step object use task (assembling and switching on a torch)’ (v) complex figure copy for constructional apraxia. Cognitive measures included both generic orientation in person, time and place, and detailed measures of Executive functions, Visuo-Spatial Attention, Tactile extinction, Memory, Language, and Calculation,. Participants were also assessed initially with the Barthel Activities of Daily Living (ADL) Index (9), a well-established and validated functional measure for acute stroke patients, which was repeated at 9 months’ follow-up. All participants gave written informed consent according to the approved Birmingham University ethics procedures.

**Table 1:** patient demographic details, stroke severity based on their ADL score and praxis results (SD: standard deviation)

|  | Initial assessment<br>(<1month) | Follow-up (at chronic<br>stage >9months) |
| --- | --- | --- |
| Number of patients | 256 | 256 |
| Avg time since stroke (**days min-max**) | 21.90 (1-90) | 285 (223-568) |
| Type of stroke | (Ischaemic 198, haemorrhagic 45,<br>Undetermined on CT 5) |  |
| Age (years +/- SD) | 69.5 (13.5) | 70.5(13.4) |
| Gender (M:F) | 146:107 | 146:107 |
| **Avg ADL score (+/-SD)** | **13.3 (5.5)** | **17.3 (3.9)** |
| Avg praxis score Gesture Production | 93 (14) | 94.4 (11) |
| Avg praxis score Meaningless Imitation | 84.84 (19) | 88.6 (14.8) |
| Avg praxis score gesture recognition | 89 (14.7) | 90.8 (13.5) |

### Statistical analyses

Statistical analyses were conducted in RStudio v.4.2.0. We conducted two sets of analyses using multivariate predictive forward stepwise regression modelling. First exploring the relationship between ADL and praxis (irrespective of time), all cognitive and praxis features were entered as predictors of ADL scores. Second, we predicted the change in ADL score between chronic and sub-acute stages using performance on cognitive and praxis measures from the sub-acute stage. All models were internally validated using 4-fold cross-validation per ‘TRIPOD’ guidelines(10).

## Results

Our time-invariant step-wise predictive modelling analyses revealed measures outlined in **Table 2** to be the most significant predictors of ADL (stepwise linear regression AIC=602.24, Rsquared = 0.30 F statistic 8.553 on 14 and 232 DF, p=7.54e-15). Three of the eight measures identified as significantly predicting ADLs (out of a total of 34 cognitive measures), related to praxis.

**Table 2:** Time invariant step wise regression results.

|  | Beta | SE | T value<br>(descending) | pValue |
| --- | --- | --- | --- | --- |
| Left Tactile Extinction | 0.032 | 0.008 | 4.05 | <6.95e-05 |
| Meaningless Gesture Imitation (Praxis) | 0.046 | 0.018 | 3.14 | 0.000747 |
| Constructional Apraxia | 0.052 | 0.0136 | 3.317 | 0.001 |
| Orientation in Time and Space | -0.053 | 0.02 | -2.6 | 0.0095 |
| Word and Non-Word Reading | 0.021 | 0.0089 | 2.37 | 0.018 |
| Gesture Production (Praxis) | -0.037 | 0.018 | -2.070 | 0.039 |
| Neglect Asymmetry | 0.022 | 0.011 | 2.19 | 0.029 |
| Neglect Total Accuracy | 0.026 | 0.012 | 2.070 | 0.039 |

Our second, step-wise linear regression model identified 60% of the variance in 9 month BI-ADL scores to be significantly explained by patients’ early subacute performance on both the ADL at baseline and on specific praxis and cognitive measures, (F(15,230)= 25.48 p-value: < 0.001, Adjusted R2: 0.59). ADL change was best predicted by ADL at early subacute stage (p < 2e-16, T= −16.146, β estimate −0.626081 SE=0.038776). Interestingly cognitive measures of limb apraxia predominantly including gesture production (or pantomime p= 0.000813, T= −3.393, β estimate=-0.055532, SE 0.016365), gesture recognition (p=0.017076, T= −2.403, β estimate: −0.034875 SE=0.014516) and meaningless gesture imitation (p= 0.004673, T= 2.857, β estimate 0.033833 SE=0.011844) were important predictors. The slope of association between each of these praxis measures and ADL change scores is displayed in **Figure 1**. Additional cognitive predictors included: orientation in time and space (p= 0.002796, T= − 3.022, β estimate −0.052508 SE=0.017376), measures of language production – including word and non-word reading (p=0.009295,T= 2.623, β estimate 0.020205, SE 0.007702), as well as sentence construction (p= 0.001568, T= −3.200, β estimate −0.040970, SE 0.012803); and finally tactile extinction to the left during bilateral stimulation (p= 0.034991, T= 2.121, β estimate 0.015937 SE= 0.007514).

**Figure 1:**
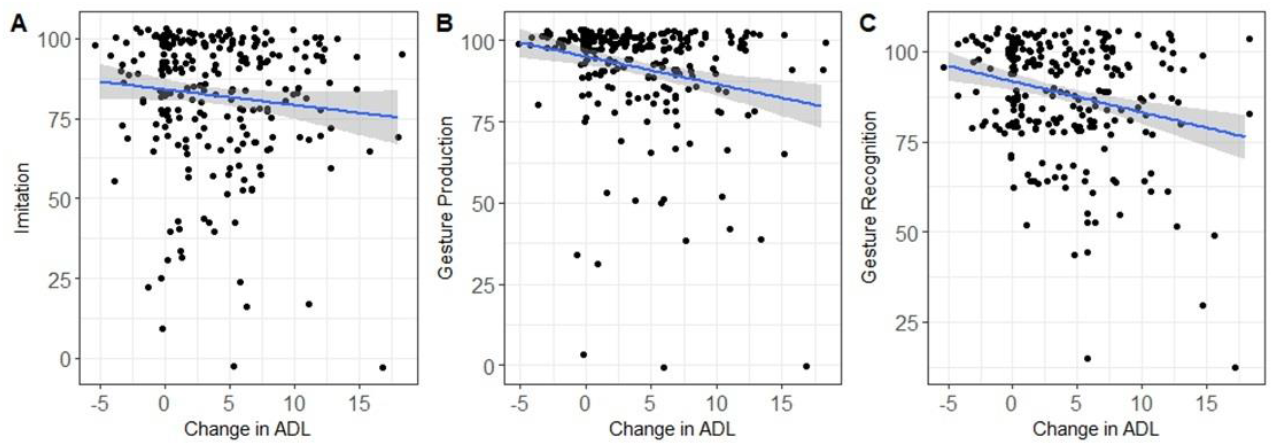

In order to assess the significance of apraxia scores in these predictions, we repeated a stepwise linear regression model *excluding* the apraxia scores and an analysis of variance comparing the model with and the one without the scores. The model without apraxia accounted for 56% of the variance (F(11,234)=29.4, pvalue <2e-16, Adjusted R2: 0.56). An ANOVA revealed a significant difference between the two models, suggesting that presence of apraxia measures outlined above was significant in predicting BI-ADL change (F=6.8, sum of square 220.92, p<3.544e-05).

## Discussion

Neurobehavioral deficits after stroke are now recognized as having a profound impact on reintegration into pre-stroke social and occupational roles, significantly limiting meaningful participation in activities of daily living (ADLs)(11). Using data from a unique longitudinal stroke patient dataset, this study provides evidence that detailed cognitive measures, mainly including apraxia measures, predict these outcomes independently.

Our results thus extend previous suggestions that praxis may be related to patient outcomes (2, 3). We showed that use of an extended screen involving several praxis and other cognitive tasks delivered at subacute stages after stroke can be predictive of patients’ ability to recover on measures of daily living activities at the chronic stage, taking into account their initial scores in daily activities. This is most important in view of the recognition of a large variability in cognitive screening tools used post-stroke in which patients’ deficits may not fully be captured. Additionally, the limitation of combining these with outcome measures has only recently been recognised. The BCOS has therefore provided us with a unique dataset comprising praxis, which is not routinely being collected when patients leave stroke units.

The Oxford Cognitive Screening (OCS) programme is the only other study that has provided a comprehensive (yet briefer) cognitive post-stroke longitudinal assessment comprising cognitive tasks along with outcome measures. However, this study only comprised one praxis task (meaningless gesture imitation) and only tested patients up to 6 months(12). They reported that apraxia based on meaningless gesture imitation is very prevalent (with 70% of patients showing deficits) in the early post-stroke stages, with a lower proportion of patients showing these deficits at 6 months, and that those who do, have greater deficits in ADLs as shown previously(3, 12). However, the ‘OCS’ study was limited by the lack of comprehensive testing for apraxia along with other cognitive measures and the fact that the ‘chronic’ post-stroke stage was only estimated at 6 months. Recovery from apraxia has been reported to take between 2 and 8 months post-stroke, with some patients recovering faster depending on the praxis tasks involved (13). Hence a follow-up of over 9 months, as the one presented here, would more accurately capture those that have recovered, thus enabling predictions about change in ADL based on praxis and cognitive measures at early subacute stages. These findings underscore the critical value of appropriate testing of cognitive functions such as that provided by BCOS in addressing the limitations of existing tools like the NIHSS and advancing our understanding of cognitive predictors of post-stroke recovery.

## Data Availability

All data produced in the present study are available upon reasonable request to the authors

## Acknowledgements

We would like to thank the patient participants taking part in the Birmingham Cognitive study as well as Professor Riddoch and the late Professor Humphreys for their original conceptualisation, and NIHR funding, which supported the BCOS study. Dr Rounis is funded by a UKRI Clinical Academic Partnership Programme, Professor Demeyere is supported by an Advanced Fellowship from the NIHR and project grant from the Stroke Association. Professor Lambon Ralph is supported by a MRC Intramural Grant.

## Author Contributions

ER, SR, and MLR contributed to conception and design of this particular study; WLB and ND in the acquisition of the original BCOS data, SR in the data analysis and preparation of the figure, ER, SR, ND and MLR in drafting the text.

## Conflicts of Interest Statement

Nothing to report.

## Notes

### Competing Interest Statement

The authors have declared no competing interest.

### Funding Statement

This study did not receive any funding

### Author Declarations

National Research Ethics Service (NRES): Essex 1 Research Ethics Committee (REC)

